# Refining Detection of Subclinical Epileptiform Activity in Alzheimer’s Disease: A Case-Control Study and Call for a Consensus

**DOI:** 10.1101/2025.10.10.25336968

**Authors:** Anna B. Szabo, Jonathan Curot, Fleur Gérard, Florence Rulquin, Rachel Debs, Claire Georges, Marie Denuelle, Amel Bouloufa, Béatrice Lemesle, Patrice Péran, Claire Thalamas, Emmanuel J. Barbeau, Jérémie Pariente, Lionel Dahan, Luc Valton

## Abstract

**Objective:** Sleep-predominant network hyperexcitability is increasingly recognized as a potential disease-accelerating comorbidity in Alzheimer’s disease (AD). However, its prevalence and risk-factors remain debated, largely due to cohort-specific and methodological differences across studies. In this prospective case-control study, we investigated potential ways of improving detection, from translational approaches focusing on REM-sleep to refined EEG setups and added clinical questionnaires.

**Methods:** We recruited 30 early-stage AD patients without a history of epilepsy and 30 age-matched controls. Participants underwent overnight polysomnography with video-EEG. Interictal epileptic discharges (IEDs) were identified through a structured three-step review by multiple independent experts using recommended criteria. Neuroanatomical patterns and sleep-related abnormalities were investigated as potential risk factors. Clinical symptoms in favor of epileptic seizures were evaluated through a tailored questionnaire at follow-up.

**Results:** IEDs were detected in three patients (10%) and one control (3.33%), a difference not reaching statistical significance (p =.612). Most events occurred during NREM sleep. Eight patients (26.67%) reported symptoms compatible with epileptic seizures - one of whom also presented with IEDs. Patients with IEDs or reported symptoms suggestive of potential seizures exhibited more severe sleep-disordered breathing and reduced precuneus volume compared to those without.

**Interpretation:** Despite efforts to optimize detection accuracy, our findings reveal a lower-than-expected percentage of AD patients with IEDs, yet support previous findings suggesting that sleep-disordered breathing and specific atrophy patterns could flag at-risk patients, guiding screening in clinical settings. Our findings also favour validation efforts of questionnaires to support the diagnostic process. Finally, we highlight methodological issues in IED detection and call for the reevaluation and standardization of diagnostic methods and criteria in this population to improve patient care.

## Introduction

An association between Alzheimer’s disease (AD) and network hyperexcitability – manifesting as interictal epileptic discharges (IEDs) and seizures, particularly during sleep^1–3^ – has been well-documented in both AD mouse models^4^ and patients^5^. Importantly, these aberrant network activities have been linked to accelerated cognitive decline across several clinical cohorts.^1,6–9^ Although their prevalence increases with disease severity^6^, IEDs have consistently been observed even in patients with mild cognitive impairment (MCI) and mild-to-moderate stage AD^1,10,11^.

Targeting network hyperexcitability using antiseizure medications, particularly levetiracetam, has shown promise. Studies report significant improvements in multiple cognitive domains among MCI and AD patients experiencing IEDs^12,13^, indicating a potential disease-modifying strategy in the face of the growing burden of AD-type dementia.

However, while IEDs are consistently detected early in nearly all AD rodent models^14^, human studies report highly variable percentages of patients with IEDs across clinical cohorts – from as low as 6%^15^ to as high as 75%^10^ – likely due to cohort-specific and methodological discrepancies^16^.

One potential avenue for better understanding this comorbidity and its discrepancies involves translational approaches that apply findings from preclinical models to human diagnostics. For example, in AD mouse models, IEDs predominantly occur during sleep and are three times more frequent during rapid eye movement (REM) sleep compared to slow-wave sleep (SWS).^17,18^ This is surprising, as REM sleep is generally considered to have an IED-suppressive effect.^19,20^ One hypothesis is that this suppressive mechanism is less effective in the mesial temporal lobe (MTL)^21^, where network hyperexcitability is thought to originate in AD.^2,22^ Nonetheless, most clinical studies do not differentiate between REM and non-REM sleep, and the characteristic EEG slowing^23,24^ or brief, unstable REM phases in AD patients may contribute to underestimating REM sleep or misclassifying IEDs as benign variants. Moreover, widespread use of antidepressants in AD^25^ may further reduce the ability to detect REM-related IEDs as these medications are known to suppress REM sleep^26,27^,.

A second promising strategy involves optimizing electrode coverage and analytic methods. Enhancing coverage of the MTL could improve IED detection^11^. Complementary qualitative tools, such as symptom-based questionnaires developed from clinical observations^16^, may also aid in identifying patients with potential underlying epileptic abnormalities^11^, helping to guide screening.

This guidance is an important point as a lack of access to examinations investigating epileptic activities is a major issue, making standard screening for all AD patients for IEDs unlikely, in spite of advocacy for this to change.^28^ Therefore, a third approach to improve diagnostic accuracy of IEDs in AD could focus on identifying high-risk subgroups among early-stage AD patients. This could be supported by tools, from the above-mentioned clinical questionnaires to flagging patients with comorbidities or distinct neuroanatomical patterns that might be linked to hyperexcitability. Although data are limited, risk factors such as more severe sleep-disordered breathing^11^ or precuneus-dominant atrophy^29^ have been proposed. However, all options mentioned above – translational approaches, electrode coverage, qualitative tools and risk-factor identification – are under-investigated and require further evaluation before clinical translation is possible.

Hence, in this paper, we present findings from a prospective observational cohort study investigating the occurrence of epileptic activity during sleep in 30 Alzheimer’s disease (AD) patients without a history of epilepsy, compared to cognitively healthy, age- and sex-matched older adults. Quantitative data on IED frequency were obtained using overnight polysomnography with concurrent video EEG (PSG-vEEG), employing extended temporal electrode coverage (25 scalp electrodes, including P9, P10, F9, F10, T9, and T10) and a rigorous three-step IED detection protocol. Qualitative data were collected through a questionnaire designed to identify symptoms suggestive of epileptic seizures.^16^

Initially, the study was designed to focus on REM-related IEDs in AD patients, building on preclinical findings that suggest a disproportionately high frequency of such discharges during REM sleep in AD models^17,18^. In parallel, and informed by prior clinical reports, we also aimed to investigate potential risk factors associated with network hyperexcitability, including sleep-disordered breathing (e.g., apneas) and structural MRI features such as precuneus-dominant atrophy. However, given the considerable variability in IED prevalence reported across studies published during the inclusion period, as well as the practical challenges encountered in accurately detecting IEDs using non-invasive methods in the current study, we also address key methodological limitations in the current diagnostic approaches and propose strategies to improve the reliability and clinical utility of IED detection in AD.

## Methodology

### Participants

The EREMAD study (NCT03923569) was a prospective, observational, single-centre case-control study conducted between April 2019 and October 2022 at the University Hospital of Toulouse and the Brain and Cognition Research Centre (France). Participants were aged 50-90 years and provided written informed consent. The study was approved by the regional ethics committee (CPP Ile de France VIII) and adhered to the Declaration of Helsinki.

The AD group comprised 31 patients with mild-to-moderate sporadic AD (MMSE ≥18), diagnosed according to IWG-2 criteria, with lumbar puncture, neuropsychological tests and, on occasions, supplementary MRIs supporting diagnosis. Note that for one patient, the diagnosis was solely based on neuropsychological assessment, overall clinical presentation, a strong family history and APOE4 homozygosity. Exclusion criteria included untreated sleep apnoea, major psychiatric/neurological illness, history of epilepsy, current use of antiseizure medication, MRI contraindications, and drugs significantly affecting sleep architecture, such as antidepressant medications or dopaminergic agonists. Neuroleptics and benzodiazepines were allowed at a maximum of one and two daily doses, respectively. None of the patients had isolated aphasia, apraxia or agnosia, sudden-onset cognitive deficits, nor non-degenerative lesions or substantial lesions in white matter with flair hyperintensities on previous MRIs. The control group included 30 cognitively normal, age- and sex-matched adults (MMSE >25, ≥9/10 on the 5-word test).

### Study Design

The protocol included two initial visits separated by no more than 60 days (V1: MRI and APOE genotyping; V2: neuropsychological assessment, overnight video-polysomnography) and one follow-up visit (V3). At V3, clinical feedback was provided, and the Epilepsy Questionnaire (EpiQ)^16^ and additional questionnaires (e.g., CRIq, fNART) were administered (**Suppl. Fig 1** and **Suppl. Table 1**). The mean interval between V1 and V3 was 336±206 days for controls and 295±184 days for patients (p=0.42). All procedures are detailed below.

### Video-EEG and IED Analysis

Overnight EEG (sampling rate: 1024 Hz) was recorded with a 27electrode setup according to the standard 10–20 setup, with extended lower temporal coverage (F9/P9/T9, F10/P10/T10). Electrodes were attached to the scalp with conductive adhesive gel (Natus) and a medical-grade tubular elastic net hood held the setup in place. A caregiver was allowed to be present throughout the recording for both groups. Recordings were manually reviewed using the DeltaMed/Natus^TM^ system in three steps (See **Suppl. Fig 2** for details) by three experts blinded to group allocation. These steps included:

Step 1: During this initial analysis carried out by two independent evaluators, the EEG recordings were scrutinized in order to detect and annotate grapho-elements suggestive of IEDs, and to conclude whether or not the recording contained potential IEDs, and if so, how many. In cases of disagreement, the grapho-elements labelled as potential IEDs were reviewed and relabelled by a third expert.

Step 2: Artifact rejection and classification of all grapho-elements that were labeled at the end of step 2 by a single rater using a five-level confidence scale: Non IED (artifacts or non-pathological variants such as Wickets spikes or Benign Sporadic Sleep Spikes (BSSS)), doubtful events, possible IEDs, probable IEDs and typical IEDs. The latter three groups were analogous to the grouping applied by Lam et al. (2020).^2^ Only possible, probable and typical IEDs were considered for the final step (see below).

Step 3: The final evaluation was based on qualitative features and quantitative criteria defined by the International Federation of Clinical Neurophysiology (IFCN).^30^ Note that the software used for the current analyses has limited ability to evaluate the sixth IFCN criterion, so decisions were based on the first five criteria and thresholds were adjusted (see **Suppl Fig. 2** for details).

### Polysomnography and sleep scoring (PSG)

The PSG-vEEG (sampled at 1024Hz) was obtained and analysed using custom-made equipment and sensors from Natus, following AASM v2.4 guidelines^31^. The setup included two disposable adhesive EOG electrodes, two chin EMG electrodes, 4 Grass® cup leg EMG electrodes, 2 ECG electrodes; thoracic and abdominal Piezo respiratory effort belts, pulse oximetry (Nonin® xPod and Nonin® PureLight® disposable adhesive sensor), a nasal airflow sensor connected via an Ultima™ Dual Pressure Flow/Snore Sensor box, a thermistor, a microphone (Ultima™) and synchronized infrared video. Sleep stages and respiratory events were scored by sleep specialists blinded to diagnostic group.

### MRI examination

The MRI took place at the Toulouse Neuroimaging Center on the site of the University Hospital in a 3T Philips ACHIEVA dStream machine (Intera Achieva, Philips, Best, The Netherlands). The current manuscript reports T1-related data acquired with the following parameters: 180 sagittal slices in 3D scan mode, multishot, voxel resolution of 1 x 1 x 1 mm³. T1-related data analysis was performed using FreeSurfer v. 7^32–34^, via a previously outlined pipeline.^35^ For further sequences used, see **Suppl. Fig. 1.**

### APO-E genotyping

APOE genotyping was performed exclusively for patients. The following kits were used, according to the manufacturer’s instructions: MagNA Pure 24 Total Nucleic Acid Kit (Roche) for nucleic acid extraction and the Fast Start DNA Master HybProbe Kit (Roche) together with the LIGHTMIX APOE C/112R R158C (TIB MOLBIOL GmbH) for the identification of APO-E haplotypes.

### Neuropsychological Assessment

As the initial study objectives also extended to investigating the impact of epileptic activities on memory consolidation, neuropsychological assessments targeting memory consolidation and general cognition were conducted before and after sleep (see **Suppl. Fig 1**). Note that given the unexpectedly uneven group distributions, and the low number of IEDs detected (see **Results**), this planned analysis had to be omitted, although descriptive results are reported in **Suppl. Table 1**.

### Follow-up visit (V3) – The EpiQ questionnaire

At follow-up, several questionnaires were completed, including the EpiQ (see **Suppl. Fig 1, Suppl. Mat. 1)**, by participants, with observation-based input from caregivers whenever possible. To assess the responses, follow-up questions were asked for positive responses to each item to elucidate the frequency, severity, and specificity of the symptoms. A decision based on these factors was made by clinical experts and participants were classified as having “potential symptoms in favor of epileptic seizures” or not.

### Main outcome variables and Statistics

Sample size was determined using a power analysis based on mean IED rates from previous studies.^1–3,15^ This indicated that with estimated effect of 5% vs. 20%, and a criterion of α = 0.05, a power of 0.85 could be achieved with 29 participants per group.

The primary outcome variables included the percentage of participants with IED on their EEG in both groups and the distribution of these IED over the sleep-wake cycle. In addition, the percentage of patients with symptoms in favor of epileptic seizure at V3 based on the EpiQ^16^ was evaluated. We also evaluated whether key structural variables from the MRI, particularly those related to temporal-lobe^36^ and the precuneus^29^, or sleep-disordered breathing-related variables differed between patients with and without IED.

Data were analysed using JASP.^37^ Group comparisons were made using t-tests, chi-square tests, Fisher’s exact test, or non-parametric equivalents as appropriate. Descriptive statistics are presented as mean (M) ± standard deviation (SD) unless otherwise noted. Due to unequal group sizes, patient group-related analyses along the IED absence/presence axis are considered exploratory and reported p-values are uncorrected. All values are rounded to two decimal places unless otherwise noted. All comparisons were two-tailed, and p-values <0.05 were considered significant.

## RESULTS

### Cohort characteristics

A total of 31 AD patients and 31 matched controls were enrolled (**Fig. 1**). After exclusion of one participant per group (due to technical failure and incidental findings), the final sample comprised 30 participants in each group. All completed the initial two visits, and all but one control attended the follow-up.

**Figure 1.**
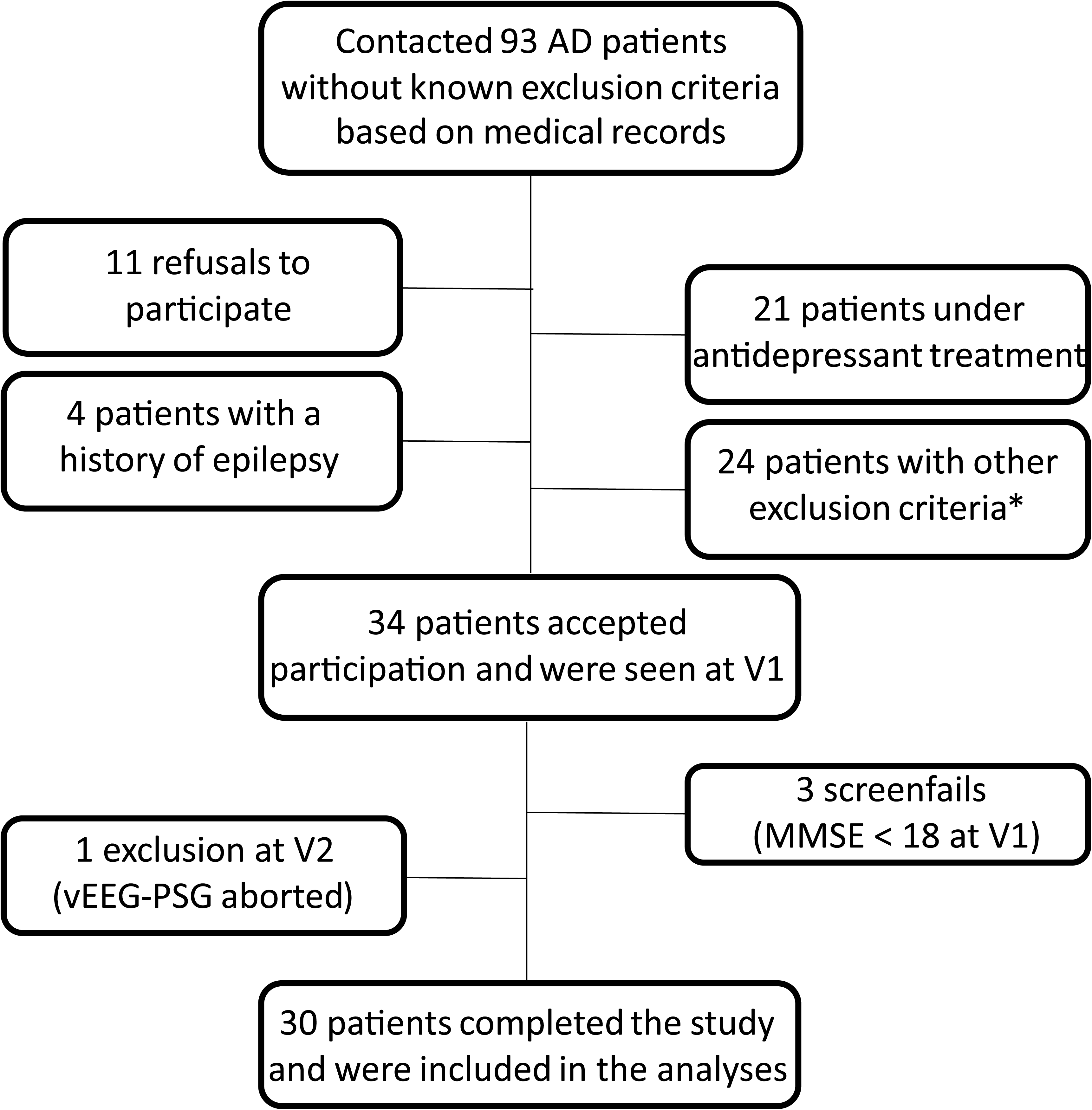
Flow chart of patient inclusions. *: Including contraindication to MRI examination, regular alcohol use, impossibility to attend the visits, neurological antecedents other than AD, participation in other clinical trials incompatible with the EREMAD study, chronic systemic pathologies, diagnosed but untreated sleep apnea syndrome, important loss of autonomy or patient already under legal guardianship/institutionalized. Abbreviations: AD = Alzheimer’s disease; MMSE = Mini-mental State Examination; vEEG-PSG = video-electroencephalogram with polysomnography.

Groups were comparable in age, sex, and key clinical variables, except for the expected MMSE difference (p < .001; Table 1). APOE ε4 carriers made up 80% of the AD group. Four AD patients (13.3%) presented with atypical variants (two logopenic, two occipital). Use of medications affecting sleep was within protocol thresholds.

**Table 1.** AD) and for patients with either IED on the EEG or an epilepsy suspicion based on EpiQ (IED+/EpiQ+ AD) and patients without IED or EpiQ-based epilepsy suspicion (IED- & EpiQ- AD). Values indicate mean (M) ± standard deviation (SD) unless stated otherwise. Reported p-values results from t-tests for the control-patient comparisons and for the IED- &EpiQ- AD vs IED+/EpiQ+ AD groups and from Mann-Whitney tests for the IED- vs IED+ patient comparison. a: n = 29 for controls. b: comparison of APOE4 heterozygotes and homozygotes. **Abbreviations** : AHT: Anti-Hypertension, CPAP: Continuous Positive Airway Pressure machine, CRIq: Cognitive Reserve Index questionnaire, MMSE: Mini Mental State Examination (Folstein Version), V1 = Visit 1, V3 = Visit Three (follow-up)

|  | Controls (n = 30) | AD patients (n = 30) | p-value Controls vs AD patients | IED- AD (n = 27) | IED+ AD (n = 3) | p-value IED- vs IED+ AD | IED/EpiQ- AD (n = 20) | IED/EpiQ+ AD (n = 10) | p-value IED/EpiQ- vs IED+/EpiQ+ AD |
| --- | --- | --- | --- | --- | --- | --- | --- | --- | --- |
| Age | 68.2 ± 5.35 | 69.67 ± 6.79 | 0.357 | 69.11 ± 6.62 | 74.67 ± 7.64 | 0.283 | 69.2 ± 6.65 | 70.6 ± 7.35 | 0.603 |
| Age at onset | NA | 68.23 ± 6.91 | NA | 67.78 ± 6.79 | 72.33 ± 8.02 | 0.387 | 68.7 ± 6.23 | 67.3 ± 8.38 | 0.609 |
| AD duration (months) | NA | 18.16 ± 27.81 | NA | 17.04 ± 28.32 | 28.47 ± 24.74 | 0.489 | 10.351 ± 20.23 | 33.86 ± 34.91 | **0.026** |
| MMSE V1 | 28.83 ± 1.05 | 24.2 ± 2.87 | **< 0.001** | 23.93 ± 2.56 | 26.67 ± 4.9 | 0.265 | 24.05 ± 2.48 | 24.5 ± 3.66 | 0.693 |
| MMSE V3, a | 28.45 ± 1.45 | 22.93 ± 4.7 | **< 0.001** | 22.56 ± 4.63 | 26.33 ± 4.73 | 0.165 | 23.6 ± 4.1 | 21.6 ± 5.72 | 0.279 |
| CRIq Education scale, a | 112.83 ± 10.92 | 115.93 ± 8.53 | 0.228 | 116.7 ± 8.66 | 109 ± 1 | 0.062 | 117.45 ± 9.13 | 112.9 ± 6.54 | 0.173 |
| Women (% (n)) | 50% (15) | 50% (15) | 1 | 48.14% (13) | 66.66% (2) | 0.543 | 45% (9) | 60% (6) | 0.439 |
| CPAP (n) | 1 | 1 | NA | 1 | 0 | NA | 1 | 0 | NA |
| Atypical AD (% (n)) | NA | 13.33% (4) | NA | 14.8% (4) | 0% (0) | NA | 5% (1) | 30% (3) | 0.058 |
| APO-E4 carriers (% (n)) | NA | 80% (24) | NA | 81.48% (22) | 66.66% (2) | 0.543 | 85% (17) | 70% (7) | 0.333 |
| APO-E4 Homozygote (% (n)), b | NA | 23.33% (7) | NA | 30.43% (7) | 0 | 0.343 | 25% (5) | 20% (2) | 0.568 |
| Statins and AHT medications | 33.33% (10) | 40% (12) | 0.592 | 37% (10) | 66.66% (2) | 0.32 | 30.43% (7) | 50% (5) | 0.429 |
| Cholinesterase inhibitor | NA | 40% (12) | NA | 40.74% (11) | 33.33% (1) | 0.804 | 40% (8) | 40% (4) | 1 |
| Benzodiazepines and nonbenzodiazepine sleep medications (% (n)) | 3.33% (1) | 6.67% (2) | 0.554 | 7.41% (2) | 0% (0) | 0.626 | 5% (1) | 10% (1) | 0.605 |

### Prevalence, morphology and topography of epileptic activities

An example of the obtained signal is shown on **Fig. 2A**. No seizures were recorded during PSG-vEEG. In the initial visual screening (Step 1), five AD patients (17%) and two controls (7%) showed graphoelements suspicious for IEDs. All annotated events from these 7 recordings (totalling 582, with 455 from AD patients, and 127 from controls) were analysed in Step 2. The majority of which were ultimately discarded as non-epileptic due to low voltage or morphology consistent with benign variants (e.g., BSSS, **Fig. 2B** or wicket spikes, **Fig. 2C**).

**Figure 2.**
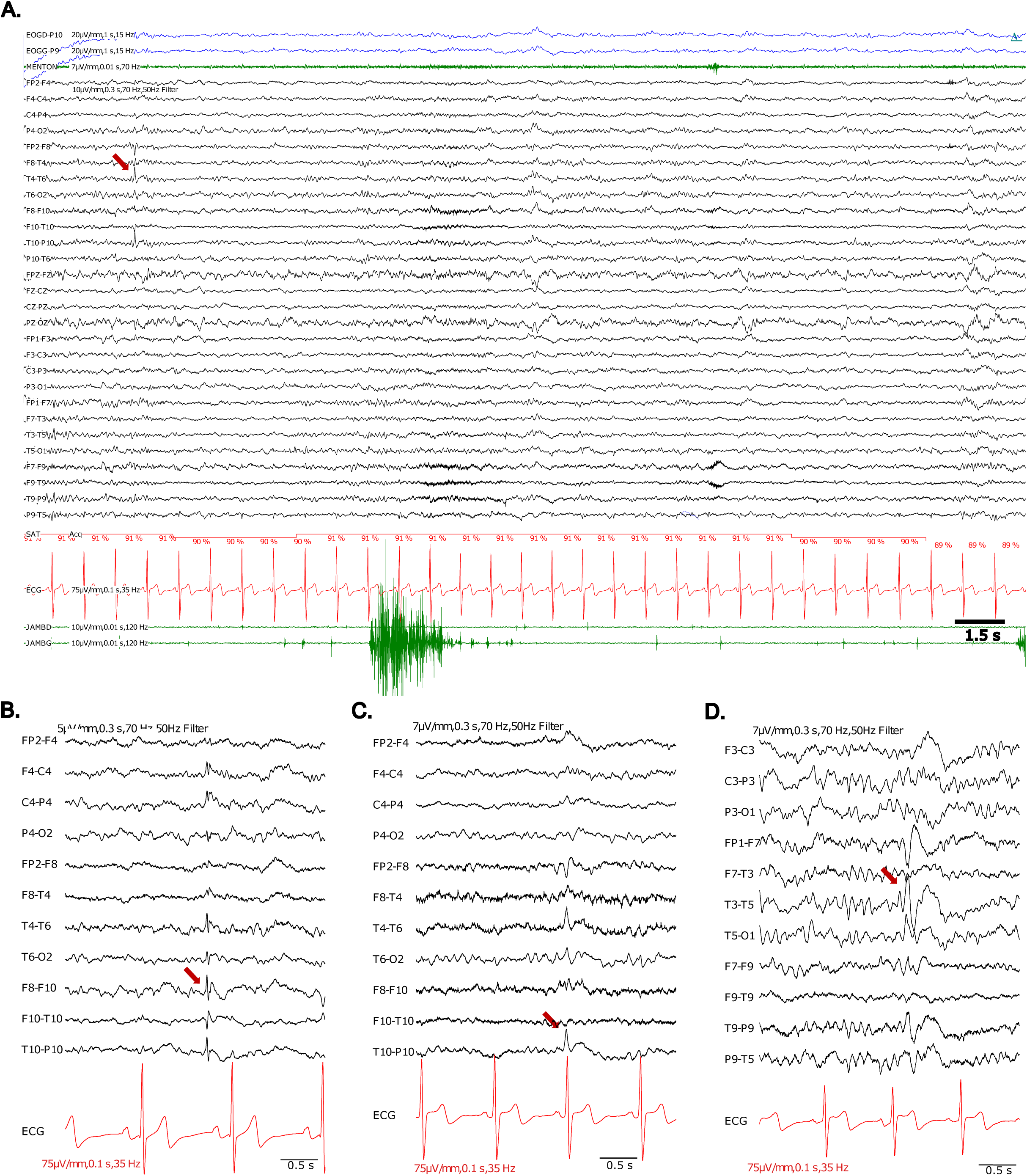
Details of the IED detection process and examples of retained IEDs and discarded graphoelements. (A) 30-second-long section of the PSG-vEEG signal with spiky graphoelement detetected at step 1 (arrow). Both sleep and epilepsy experts were free to modify the view (gain, channels shown, etc.). For clarity, breathing-related channels were removed from the illustration. (B) Graphoelement discarded as a benign variant, (BSSS). (C) IED-like graphoelement with a temporal synchrony with ECG activity. This sharp wave was initially qualified as a possible IED (step 1), and was requalified during step 2, as a non-pathological variants (probable wicket spike) and excluded. (D) Retained IED. For (B-D), the segments are ∼3.5 seconds long, with scales shown in the bottom right for time and on the top (EEG) or bottom (ECG) left for gain.

In Step 3, 24 events from four AD patients (13%) and 21 from one control (3%) were reviewed against IFCN criteria. Events meeting at least two criteria were classified as confirmed IEDs. Ultimately, IEDs were confirmed in three AD patients (10%) and one control (3.3%), with nine IEDs retained per group. Most events were temporal in location, mostly unilateral in AD and bilateral in the control with IED and consisted primarily of simple spikes (**Fig. 2D**).

The difference in IED prevalence between groups was not statistically significant (OR = 3.22; 95% CI: 0.32–32.89; χ² = 1.071; p = 0.30).

The observed comparability of REM sleep percentages between patients and controls (**Fig. 3A**) suggests that removing antidepressants from the study indeed increased REM percentages and allowed for sufficient REM to detect eventual IEDs in this stage. Furthermore, no differences in REM sleep percentages were observed when comparing IED+ and IED-patients (**Fig. 3B**), suggesting that the lack of REM sleep would not be a reason for missing out on REM-related IEDs. Yet, the majority of detected IEDs occurred during NREM2 or other NREM stages and never during REM (**Fig. 3C**), suggesting that the observed IED pattern in AD models does not translate to clinical populations.

**Figure 3.**
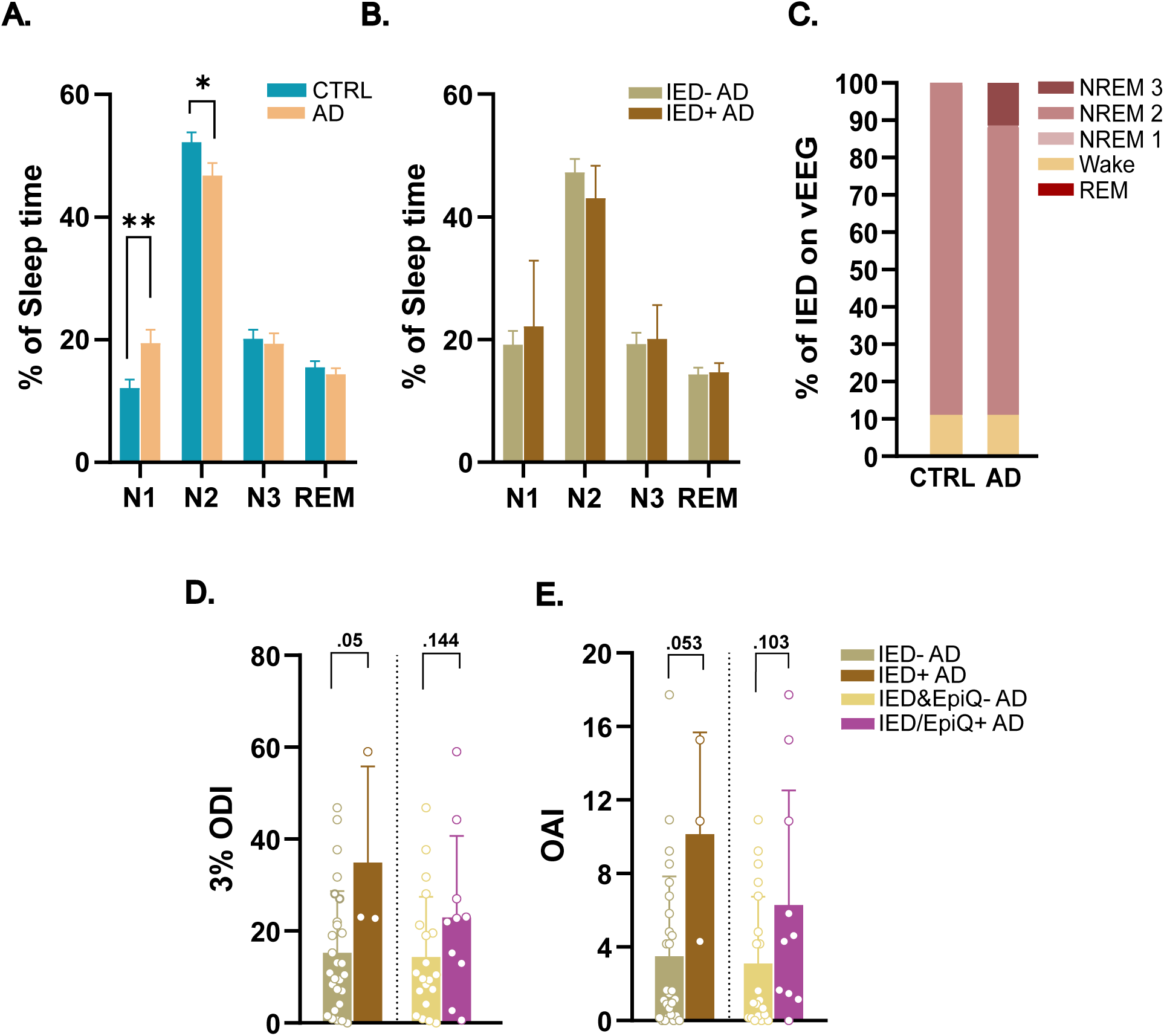
Comparison of percentage spent in different sleep stages between **(A)** all controls and patients and **(B)** between patients with or without IEDs on their EEGs, showing no significant difference between REM percentages, indicating that any observed differences for IEDs in this stage could not be attributed to REM differences. **(C)** The distribution of IED over the sleep-wake cycle across patients and controls, showing no REM-related IEDs. **(D)** 3% Oxygen Desaturation Index and **(E)** Obstructive Apnea Index, indicating a slight increase of sleep-disordered breathing in patients with IED compared to those without. Color codes for panels D and E are identical. Bars and error bars on each panel represent Mean+SEM. Statistics result from t-tests. Abbreviations: ODI: Oxygen desaturation index. OAI: Obstructive Apnea Index.

### Questionnaire-Reported Seizure Symptoms (EpiQ)

No patient nor control has been hospitalised with or treated for epileptic seizures, during the follow up period. According to the EpiQ, 8 of 30 AD patients (27%) but no controls reported symptoms indicative of possible epileptic seizures, and these symptoms led to initiate anti-seizure medication (ASM), at the follow-up visit, in two of these 8 patients. Only one of these eight patients had confirmed IEDs; the remaining showed either unconfirmed graphoelements or none.

Of note, EpiQ+ symptoms were more common in patients with any suspicious EEG features in Step 1 (4/7; 57%) than in those without (4/23; 17%; p = 0.037), suggesting that current morphological criteria may exclude true epileptiform activity in this patient population.

### Risk factors associated with IED in AD

Based on the results of the questionnaire, and to better capture the risk factors of subclinical epileptic activity, we analysed two sets of contrasts: (1) Confirmed IEDs (IED+; n = 3) vs. No IEDs (IED−; n = 27) and (2) Patients with either IEDs or EpiQ+ symptoms (IED/EpiQ+; n = 10) vs. Neither (IED&EpiQ−; n = 20). Note that both of these analyses are considered exploratory due to the unequal sample sizes.

No significant differences in age, MMSE, or disease duration were found between IED+ and IED-patients. However, IED/EpiQ+ patients had longer disease duration (M = 33.9 vs. 10.4 months, p = 0.026) and a trend toward more atypical AD variants (30% vs. 5%, p = 0.058) than IED&EpiQ-patients, although the low proportion of atypical patients in the cohort warrants caution with interpretation.

However, in line with a previous report suggesting higher IED prevalence in AD patients with sleep-disordered breathing, we observed an increased 3% oxygen desaturation index (3% ODI) in the IED+ group compared to the IED-group (23±18 vs. 11±16, p=0.05, **Fig. 3D**). This is likely due to the tendency of IED+ patients to have more obstructive apneas (Obstructive Apnea Index: 11±5 vs. 1±5, p=0.053, **Table 2**, **Fig. 3E**). Furthermore, all patients in the IED+ group received a diagnosis of moderate (n=2) or severe (n=1) sleep apnea syndrome. While the same comparisons were not significant between IED/EpiQ+ vs IED&EpiQ-patients (**Fig. 3E**), moderate-to-severe OSA was diagnosed in almost all IED/EpiQ+ patients (90%) and only about half of the IED&EpiQ-patients (55%) (p = 0.055), which may point towards more severe sleep-disordered breathing in this group.

**Table 2.** Table of sleep-related variables between different groups. Values indicate mean ± standard deviation unless stated otherwise. Reported p-values results from t-tests for control-AD comparisons (unless otherwise indicated) and from Mann-Whitney tests for the IED-/IED+ AD comparison, and, given a high number of compared variable with failed assumption checks, for the IED-& EpiQ- vs IED+/EpiQ+ AD comparisons as well. **Abbreviations** : NREM: Non rapid-eye movement sleep, OAI: Obstructive Apnea Index, ODI: Oxygen Desaturation Index, SAS: Sleep Apnea Syndrome a: Mann-Whitney U test due to violated assumptions of the Student T-Test. b: n=29/group, as participants already equipped were not considered, c: Chi-Squared test

|  | Controls (n = 30) | AD patients (n = 30) | p-value<br>Controls vs<br>AD patients | IED- AD (n = 27) | IED+ AD (n = 3) | p-value IED-<br>vs IED+ AD | IED/EpiQ- AD<br>(n = 20) | IED/EpiQ+ AD<br>(n = 10) | p-value<br>IED/EpiQ- vs<br>IED+/EpiQ+<br>AD |
| --- | --- | --- | --- | --- | --- | --- | --- | --- | --- |
| NREM 1% | 12.09 ± 7.85 (a) | 19.46 ± 11.98 | 0.017 | 19.17±11.51 | 22.14±18.56 | 0.845 | 18.64±12.2 | 21.11±11.99 | 0.559 |
| NREM 2% | 52.21 ± 8.85 | 46.8 ± 11.26 | 0.043 | 47.22±11.53 | 43.04±9.16 | 0.509 | 47.59±12.11 | 45.22±9.71 | 0.619 |
| NREM 3% | 20.16 ± 8.22 | 19.35 ± 9.36 | 0.725 | 19.26±9.53 | 20.12±9.5 | 0.948 | 18.83±8.56 | 20.4±11.23 | 0.650 |
| REM % | 15.53 ± 5.51 | 14.36 ± 5.37 | 0.412 | 14.33±5.62 | 14.68±2.59 | 0.897 | 14.92±5.64 | 13.25±4.87 | 0.350 |
| OAI | 2.8 ± 4.26 | 4.17 ± 4.8 | 0.245 | 3.51±4.33 | 10.15±5.53 | 0.053 | 3.12±3.63 | 6.29±6.24 | 0.103 |
| AHI | 13.36 ± 14.57 | 16.58 ± 13.46 | 0.378 | 15.42±12.52 | 27±20.04 | 0.3 | 20.08±15.25 | 28.58±15.04 | 0.118 |
| 3% ODI | 13.34 ± 12.91 | 17.24 ± 15.07 | 0.287 | 15.27±13.4 | 34.93±20.89 | **0.050** | 14.37±13.08 | 22.96±17.77 | 0.143 |
| Novel moderate-to-severe SAS diagnosis (c) | 46.67% (14) | 66.67% (20) | 0.12 | 62.96% (17) | 100% (3) | N/A | 55% (11) | 90% (9) | 0.056 |

On the MRI, IED/EpiQ+ patients had significantly lower total and left precuneus volume and thickness (p = 0.019–0.026), with a similar trend on the right (p = 0.06). Whole left-hemisphere cortical volume was also reduced (p = 0.033, **Table 3**). Hippocampus-related metrics were not significantly different across groups.

**Table 3.**
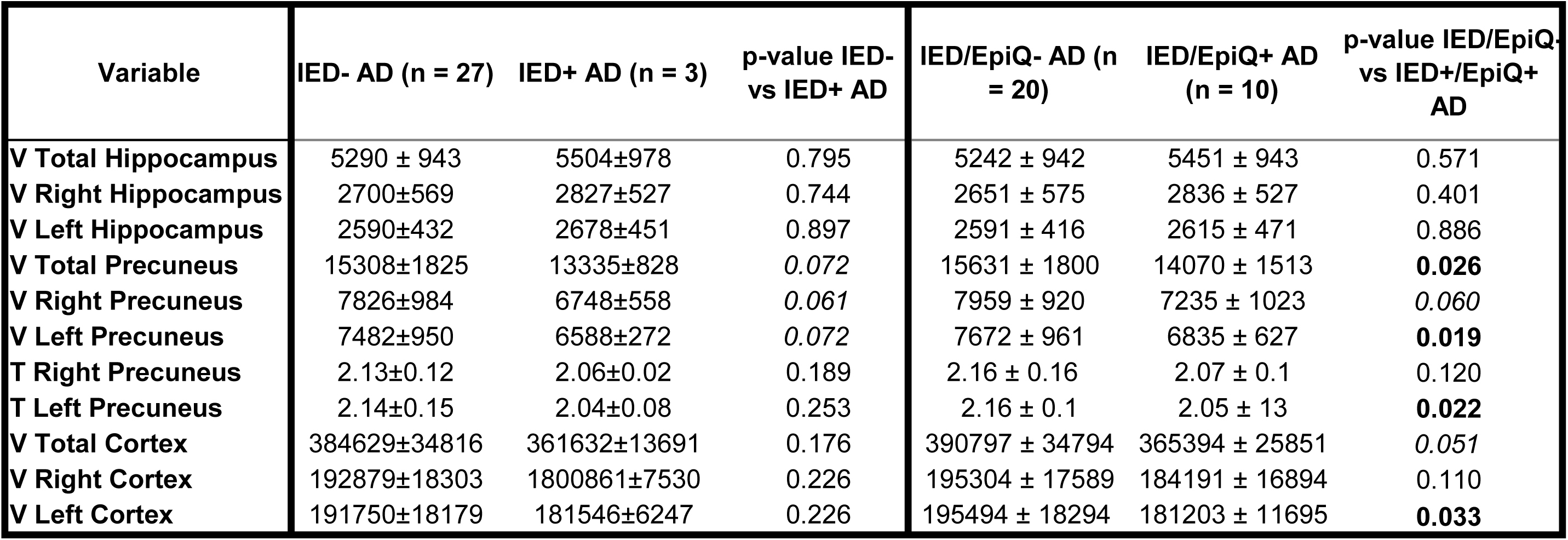
Table of morphometric variables for patients with (IED+ AD) or without IED on their EEG (IED-AD) and for patients with either IED on the EEG or an epilepsy suspicion based on EpiQ (IED/EpiQ+ AD) and patients without IED or EpiQ-based epilepsy suspicion (IED& EpiQ-AD). Reported p-values results from t-tests for the IED-&EpiQ-AD vs IED+/EpiQ+ AD groups and from Mann-Whitney tests for the IED- vs IED+ patient comparison. Values indicate Mean ± SD rounded to two decimals for thickness and to integers for volume values, unless stated otherwise. Volumes are calculated in mm3, thickness in mm. Abbreviations: T: Thickness V: Volume.

| Variable | IED- AD (n = 27) | IED+ AD (n = 3) | p-value IED- vs IED+ AD | IED/EpiQ- AD (n = 20) | IED/EpiQ+ AD (n = 10) | p-value IED/EpiQ- vs IED+/EpiQ+ AD |
| --- | --- | --- | --- | --- | --- | --- |
| V Total Hippocampus | 5290 ± 943 | 5504±978 | 0.795 | 5242 ± 942 | 5451 ± 943 | 0.571 |
| V Right Hippocampus | 2700±569 | 2827±527 | 0.744 | 2651 ± 575 | 2836 ± 527 | 0.401 |
| V Left Hippocampus | 2590±432 | 2678±451 | 0.897 | 2591 ± 416 | 2615 ± 471 | 0.886 |
| V Total Precuneus | 15308±1825 | 13335±828 | 0.072 | 15631 ± 1800 | 14070 ± 1513 | **0.026** |
| V Right Precuneus | 7826±984 | 6748±558 | 0.061 | 7959 ± 920 | 7235 ± 1023 | 0.060 |
| V Left Precuneus | 7482±950 | 6588±272 | 0.072 | 7672 ± 961 | 6835 ± 627 | **0.019** |
| T Right Precuneus | 2.13±0.12 | 2.06±0.02 | 0.189 | 2.16 ± 0.16 | 2.07 ± 0.1 | 0.120 |
| T Left Precuneus | 2.14±0.15 | 2.04±0.08 | 0.253 | 2.16 ± 0.1 | 2.05 ± 13 | **0.022** |
| V Total Cortex | 384629±34816 | 361632±13691 | 0.176 | 390797 ± 34794 | 365394 ± 25851 | 0.051 |
| V Right Cortex | 192879±18303 | 1800861±7530 | 0.226 | 195304 ± 17589 | 184191 ± 16894 | 0.110 |
| V Left Cortex | 191750±18179 | 181546±6247 | 0.226 | 195494 ± 18294 | 181203 ± 11695 | **0.033** |

## DISCUSSION

In this prospective case-control study, we deployed a full-night vEEG-PSG with an electrode setup extended with lower temporal electrodes to maximize our chances to detect IEDs during sleep in AD. Using a rigorous method for IED identification, only 10% of the patients and 3% of the controls exhibited IEDs. In patients, IEDs were mainly detected on temporal electrodes during NREM2 sleep but, contrary to our hypothesis, never during REM sleep. The EpiQ questionnaire allowed identifying eight AD patients who presented symptoms in favor of epileptic seizure, one of whom had confirmed IEDs. Although further validation is needed, our findings align with prior reports suggesting an association between epileptic activity and both sleep-disordered breathing and reduced precuneus volume^11,29^.

### IED frequency and distribution over sleep stages

Our findings fall at the lower end of the wide range of IED prevalence reported in AD (6– 75%).^1–3,6,11,15^ Furthermore, in contrast to AD mouse models overexpressing mutated APP – where REM sleep is associated with increased IEDs – we detected no IED during REM, despite efforts to preserve REM architecture by excluding REM-suppressing medications. Instead, similar to patterns seen in non-AD-related MTL epilepsy, IEDs occurred predominantly during NREM2 sleep, when hippocampal synchrony is heightened.^38^

### Cohort-specific factors behind discrepant results

Variability across studies likely reflects cohort and protocol differences. Our AD group had a relatively high average MMSE (24), possibly contributing to the low IED prevalence, as epileptic activity increases with disease severity.^5,39^ The exclusion of patients on antidepressants may also have played a role. Although these drugs reduce REM sleep, they may also increase seizure susceptibility in temporal lobe epilepsy^40,41^, while depression may increase the risk of epilepsy in itself.^42^ One report also suggests that fluoxetine triggers seizures in an AD mouse model.^43^ Interestingly, Brunetti et al.^15^, who excluded patients on psychoactive medications including antidepressants, reported similarly low IED prevalence (6.38% in AD and 11.63% in MCI patients). However, two other studies that included antidepressant users and reported on related group differences did not observe a significant effect.^1,6^ Given the growing use of antidepressants in AD^44^, further investigation is warranted regarding this point.

### Methodology related factors behind discrepant results

The method used to detect IED on the EEG is of central interest when examining discrepant results. In particular, standard clinical tools, such as scalp EEG using the 10–20 setup and single-night recordings, have limited sensitivity. For instance, single-night EEG captured IEDs in only approximately 50% of AD patients with known epilepsy^2^. Similarly, combined intracranial and scalp EEG data from two AD patients showed that merely 5% of events recorded intracranially were visible on scalp EEG^45^. This would imply that (1) the 3 patients with 2-4 IEDs detected in our study could have, in reality, a hundred throughout the night and that (2) we may not have been able to detect any IEDs in some patients.

However, while sEEG or other alternatives, for example magnetoencephalography (MEG), may offer higher sensitivity^1,6,46^, their use is restricted by ethical, logistical, or financial limitations. While emerging technologies, such as optically pumped magnetometer-based MEG and advanced computational analysis tools^47–49^, may offer more viable diagnostic solutions in the future, the current clinical reality calls for improved non-invasive strategies to enhance the detection of epileptic activity in AD and MCI. This could facilitate earlier intervention and clinically meaningful cognitive improvements.

Within the current clinical context and array of diagnostic options, we used lower temporal electrodes to improve detection accuracy – now supported by another study^11^ – and concomitant PSG recording to eliminate artifacts. While the reported IED-occurrence is not in favour of improved detection – as we hypothesised – the added lower temporal electrodes proved to be a valuable addition to the setup, based on the subjective evaluation of the experts. Nevertheless, our use of IFCN criteria to minimize false positives may have excluded genuine IEDs due to their atypical morphology in AD, which remains to be clarified. For example, based on the morphology of the examples presented in Lam et al. (2020), using our methodology would likely have led us to retain only what they called “definite” IED and to discard IED classified as “probable”, or “equivocal” which accounted for 59% of the IED in that study.

The EpiQ questionnaire added a subjective clinical dimension. Although only one EpiQ+ patient had confirmed IEDs, many patients with suspicious but unconfirmed graphoelements also reported relevant symptoms, echoing findings via a similar clinical tool.^11^ This may suggest that AD-related IEDs may differ morphologically from typical mesial temporal lobe epilepsy and may not always align with existing criteria. Some rejected elements (e.g., >400 BSSS in one patient) might in fact reflect pathological activity. In light of potential morphological differences and differences in IED definitions across studies, consensus on IED morphology and detection methods specific to AD – based on expert review and machine learning approaches – is urgently needed if we are to better understand this comorbidity. Until then, questionnaires like EpiQ may complement EEG in identifying network hyperexcitability and merit further validation in larger cohorts.

### Potential risk factors of IEDs in AD warranting screening

EpiQ could, therefore, be a useful questionnaire to identify AD patients who might require EEG screening for IEDs – although once again, this would require better screening tools. Consensus based on accumulated data from studies published so far could help identify other useful biomarkers and standardize their use in this cohort.

Nonetheless, the fact that two out of three patients with confirmed IEDs reported no symptoms in favor of possible epileptic seizures may call for the identification of further risk factors that might guide screening. According to our results and in light with previous reports^11,50^, this could include precuneus-dominant atrophy patterns and more severe forms of obstructive sleep apnoeas. While results are still based on few patients and require further studies, especially for understanding the direction of the links, such relationships are mechanistically plausible. Indeed, OSA is linked to increased amyloid burden in older adults in the hippocampus and the precuneus^51,52^, and these areas have been identified as the primary loci of network hyperexcitability in AD models or patients^53,54^. Interestingly, a recent hypothesis suggest an initial role of Aβ accumulation in the posteromedial cortex (PMC, including the precuneus) to the dysconnectivity between the MTL and the PMC, that would drive hyperexcitability in the MTL in AD.^55^ In this context, it is possible that over time, undetected OSA accelerates hippocampal and precuneal Aβ generation, increasing the risk of AD and exacerbating Aβ-induced neuronal hyperexcitability. While OSA may be a notable contributor, its treatment, even in mild-to-moderate AD, may face challenges related to CPAP tolerance. This pledges toward better screening for OSA in middle-aged and older adults to initiate treatment before cognitive difficulties impede treatment tolerance and to explore alternative treatment options such as mandibular advancement devices or, when appropriate, positional therapy.

## Conclusions

In summary, we attempted to optimize IED detection in AD through enhanced EEG electrode coverage, rigorous diagnostic criteria, and the addition of a questionnaire aimed at capturing symptoms suggestive of underlying epileptic seizures. We also tested, in a translational approach, whether REM sleep could be a target for improved IED detection. Still, only 10% of AD patients exhibited IED, and these occurred preferentially during NREM2. In spite of the low IED rate, our study brings important methodological insights, and suggests in accordance with previous studies, potential for improving screening through clinical questionnaires or through flagging patients at risk for IEDs based on comorbidities such as moderate-to-severe OSA or a precuneus-dominant atrophy pattern.

Detection of subclinical epileptic activities in AD remains a significant challenge. Future progress will depend on developing more sensitive automated IED detection tools^47,48^ and validating tailored clinical screening instruments. While our study had several limitations – many brought up previously, such as the use of scalp EEG, or the limited statistical power in subgroup comparisons, and the necessity to validate the EpiQ questionnaire – it adds valuable insights to the field regarding potential underlying factors of discrepancies between clinical studies, and calls for the reevaluation and standardization of IED detection methods for MCI and AD patients. Given that treatment of subclinical epileptic events may improve cognition in both AD and MCI^1,13^, optimizing detection remains a clinical priority.

## Abbreviations

AD: Alzheimer’s disease
BSSS: Benign Sporadic Sleep Spikes
IEDs: interictal epileptic discharges
MTL: Mesial Temporal Lobe
(N)REM: (non)-rapid eye movement
OSA: obstructive sleep apnea
PSG-vEEG: polysomnography with video-EEG
APO-E: apolipoptorein E
MMSE: mini-mental state examination
EEG: Electroencephalography

## Acknowledgements and Funding

The authors thank Brigitte Pouzet, Sandrine Rolland and all members of the Clinical Investigation Center of the CHU Toulouse for participant recruitment and administrative work. We are grateful to the nurses of the Sleep and Epilepsy Unit of the CHU Toulouse and the members of the Toulouse Neuroimaging Center for polysomnography and MRI acquisitions, respectively. The authors thank Virginie Jacques of the Federal Institute of Biology, CHU Toulouse - Laboratory of Molecular Biology and Constitutional Genetics for the APOE analyses. Finally, we thank all patients and controls for their valuable participation.

The association *Fondation Alzheimer* (France) funded the study. A. B. Szabo received PhD scholarship from the association *France Alzheimer*. F. Gérard received residency-research funding from the *Regional Health Agency of the Occitanie Region* (ARS Occitanie, France). A. Bouloufa received MSc internship funding from the *Toulouse Mind and Brain Institute* (TMBI, France). The authors are grateful for these organizations having made this clinical study possible as well as to the CHU Toulouse for being the promoter of the study.

## Author Contributions

**Conception and funding acquisition**: LD, LV, EB, JC, RD, CT, JP, ABS; **Data Acquisition**: ABS, FG; **EEG analysis:** LV, JC, MD; **PSG analysis:** RD, JC, CG; **MRI analysis**: PP; **Statistical analysis**: ABS, AB; **Manuscript drafting and figure preparation**: ABS; **Scientific advisor:** LD; **Principal investigator:** LV. All authors contributed to the revision of the manuscript and accepted the final version.

## Data Availability Statement

Data are available in the supplemental tables. All PSG raw data are available upon request by email to, except for video data, which have been deleted to conform to the ethics committee’s recommendations.

---

**Suppl. Fig. 1.**
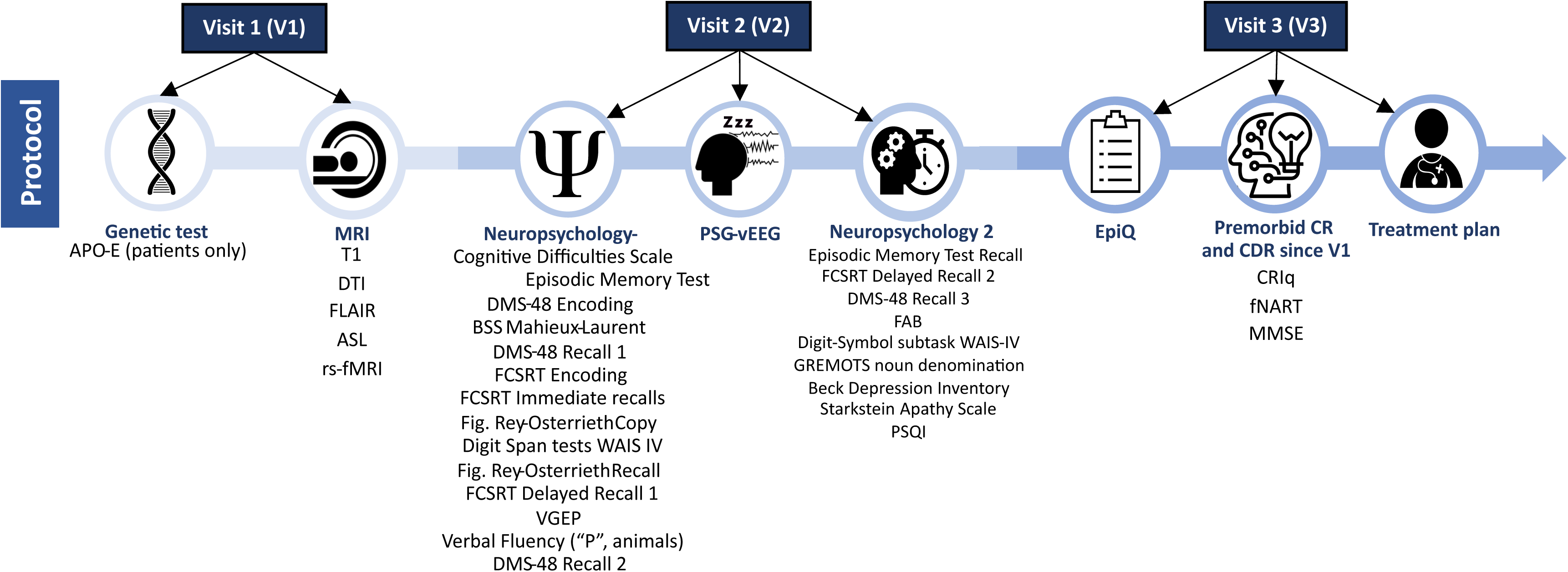
The experimental protocol, divided into 3 visits, describing the specific examinations taking place at each visit.

**Suppl. Fig. 2.**
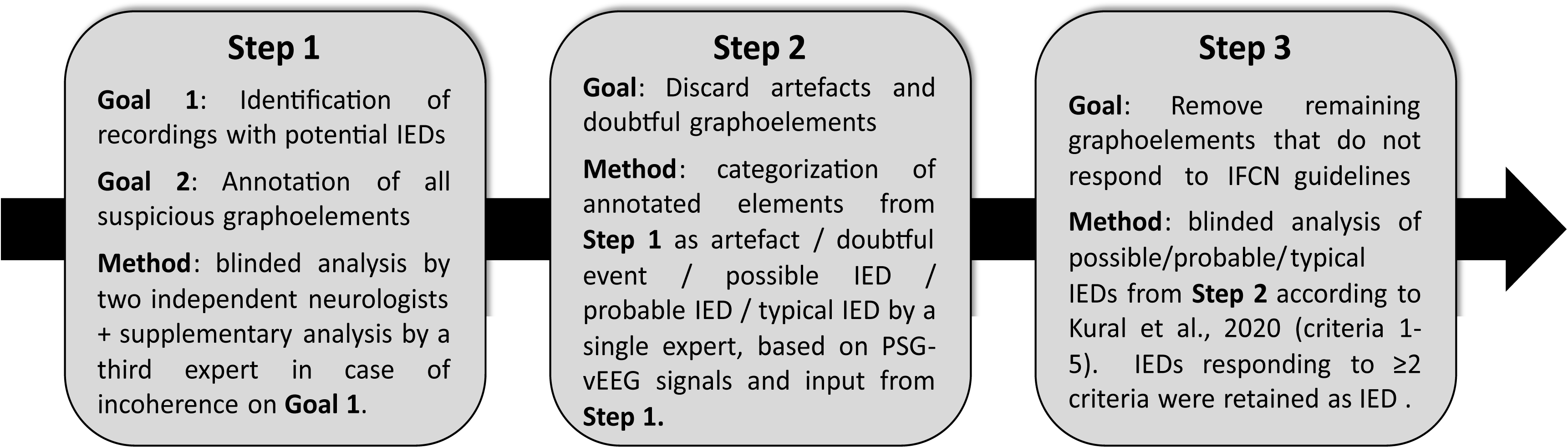
Steps included in the manual IED identification pipeline.

**Supplementary table 1.**
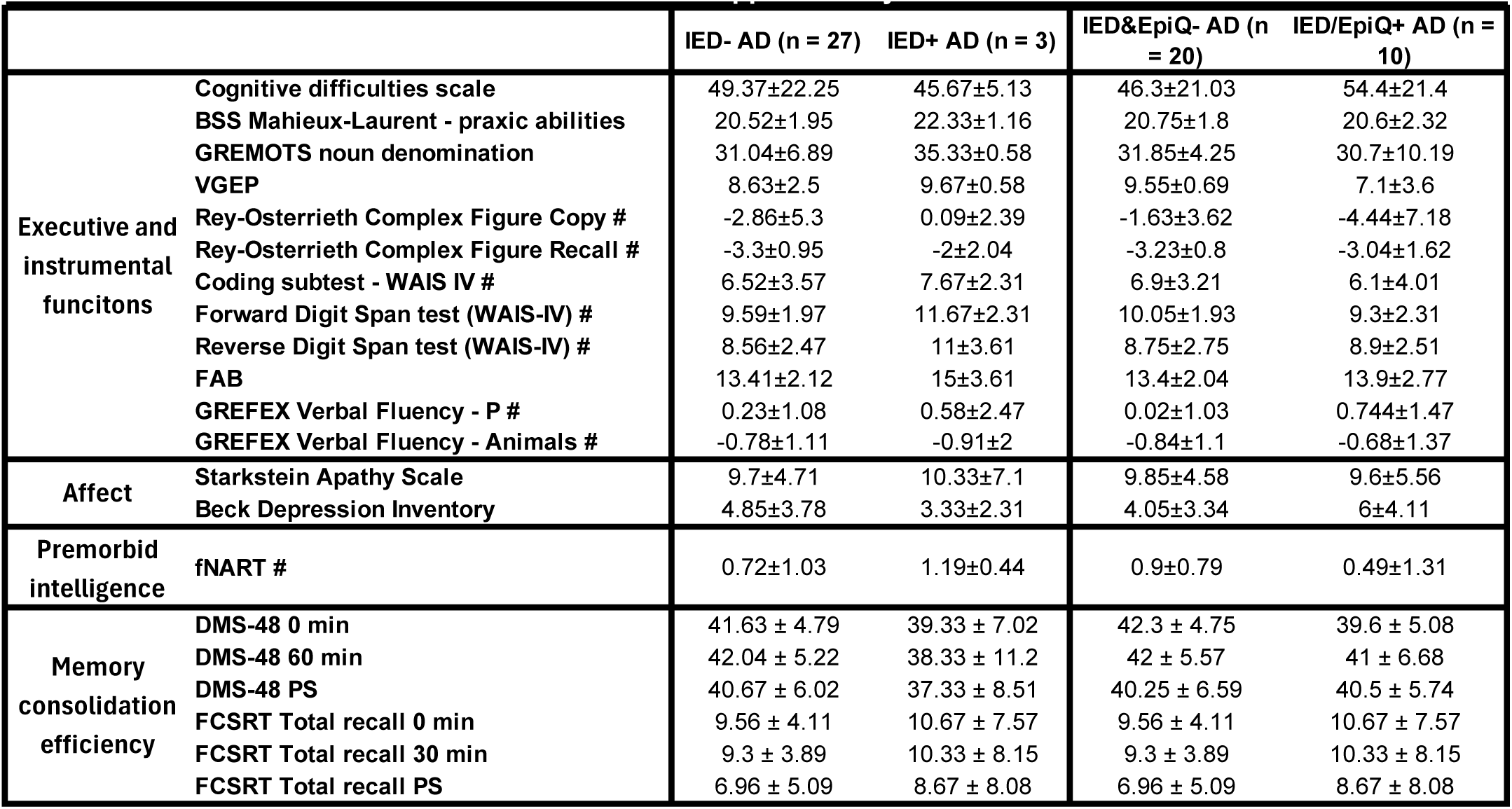
Descriptive results of neuropsychology-related variables from different patient groupings (IED+ and IED-, and IED&EpiQ- and IED/EpiQ+). Values indicate mean ± standard deviation rounded up to two decimals, unless stated otherwise. Tests where participants did not respond were scored as 0. Results reported on tests marked with # are standardized scores. **Abbreviations** : BSS: Brief Screening Scale, VGEP: Visual Gnosis Evaluation Protocol; fNART: National Adult Reading Test, French version (scores standardized for the presence of dementia).

## Supplementary Material 1 - Epilepsy questionnaire

**<u>Instructions</u>:** This questionnaire is intended to explore the occurrence of symptoms indicative of prior epileptic seizures. Given the frequent losses of consciousness or amnestic post-ictal episodes, it is preferred for the interview to be conducted in the presence of a caregiver or a person living with the patient.

|  |  |  |  |  |
| --- | --- | --- | --- | --- |
| **Presence of caregiver:** | Yes ☐ |  | No ☐ |  |
|  | ACCORDING TO THE PATIENT |  | ACCORDING TO CAREGIVER<br>(IF APPLICABLE) |  |
|  | <u>YES</u> | <u>NO</u> | <u>YES</u> | <u>NO</u> |
| <u>1-</u> SENSORY DISTURBANCE (UNPROVOKED PAIN, TINGLING, PRICKING, NUMB SKIN) | ☐ | ☐ | ☐ | ☐ |
| <u>2-</u> UNEXPLAINED SENSATION OF FEAR | ☐ | ☐ | ☐ | ☐ |
| <u>3-</u> TRANSITORY MEMORY IMPAIRMENT | ☐ | ☐ | ☐ | ☐ |
| <u>4-</u> OLFACTORY HALLUCINATIONS | ☐ | ☐ | ☐ | ☐ |
| <u>5-</u> DEJA-VU | ☐ | ☐ | ☐ | ☐ |
| <u>6-</u> TRANSIENT MOTOR DISTURBANCE (LOSS OF MUSCLE TONE, PARESIS) | ☐ | ☐ | ☐ | ☐ |
| <u>7-</u> TRANSITORY LANGUAGE IMPAIRMENT | ☐ | ☐ | ☐ | ☐ |
| <u>8-</u> FEELING OF ASCENDING EPIGASTRIC WARMTH | ☐ | ☐ | ☐ | ☐ |
| <u>9-</u> OROFACIAL AUTOMATISMS | ☐ | ☐ | ☐ | ☐ |
| <u>10-</u> MOTOR AUTOMATISMS (E.G. RHYTHMIC OR STEREOTYPED MOVEMENTS) | ☐ | ☐ | ☐ | ☐ |
| <u>11-</u> STRONG VERTIGO | ☐ | ☐ | ☐ | ☐ |
| <u>12-</u> AUDITORY HALLUCINATIONS | ☐ | ☐ | ☐ | ☐ |
| <u>13-</u> SUDDEN ONSET OF CONFUSION | ☐ | ☐ | ☐ | ☐ |
| <u>14-</u> ABNORMAL MOUVEMENTS (MUSCLE TWITCHES/MYOCLONUS) | ☐ | ☐ | ☐ | ☐ |
| <u>15-</u> LOSS OF CONSCIOUSNESS | ☐ | ☐ | ☐ | ☐ |
| <u>16-</u> CONVULSIVE SEIZURES | ☐ | ☐ | ☐ | ☐ |
| <u>17-</u> AUTONOMIC SENSATIONS (TACHYCARDY, TRANSPIRATION, | ☐ | ☐ | ☐ | ☐ |
| <u>18-</u> URINARY LEAKING AT AWAKENING | ☐ | ☐ | ☐ | ☐ |
| <u>19-</u> TONGUE BITING AT AWAKENING | ☐ | ☐ | ☐ | ☐ |
| <u>20-</u> TRANSITORY ALTERED STATE OF MIND, LAPSES IN AWARENESS | ☐ | ☐ | ☐ | ☐ |

## References

1. Vossel KA, Ranasinghe KG, Beagle AJ, et al. Incidence and impact of subclinical epileptiform activity in Alzheimer’s disease. Ann. Neurol. 2016;80(6):858–870.

2. Lam AD, Sarkis RA, Pellerin KR, et al. Association of epileptiform abnormalities and seizures in Alzheimer disease [Internet]. Neurology 2020;95(16)[cited 2024 Apr 17 ] Available from: https://www.neurology.org/doi/10.1212/WNL.0000000000010612

3. Horváth A, Szűcs A, Hidasi Z, et al. Prevalence, Semiology, and Risk Factors of Epilepsy in Alzheimer’s Disease: An Ambulatory EEG Study. J. Alzheimer’s Dis. 2018;63(3):1045–1054.

4. Palop JJ, Chin J, Roberson ED, et al. Aberrant Excitatory Neuronal Activity and Compensatory Remodeling of Inhibitory Hippocampal Circuits in Mouse Models of Alzheimer’s Disease. Neuron 2007;55(5):697–711.

5. Amatniek JC, Hauser WA, DelCastillo-Castaneda C, et al. Incidence and Predictors of Seizures in Patients with Alzheimer’s Disease. Epilepsia 2006;47(5):867–872.

6. Nous A, Seynaeve L, Feys O, et al. Subclinical epileptiform activity in the Alzheimer continuum: association with disease, cognition and detection method. Alzheimers Res. Ther. 2024;16(1):19.

7. Horvath AA, Papp A, Zsuffa J, et al. Subclinical epileptiform activity accelerates the progression of Alzheimer’s disease: A long-term EEG study. Clin. Neurophysiol. 2021;132(8):1982–1989.

8. Vöglein J, Ricard I, Noachtar S, et al. Seizures in Alzheimer’s disease are highly recurrent and associated with a poor disease course. J. Neurol. 2020;267(10):2941–2948.

9. Haoudy S, Jonveaux T, Puisieux S, et al. Epilepsy in Early Onset Alzheimer’s Disease. J. Alzheimer’s Dis. 2022;85(2):615–626.

10. Musaeus CS, Frederiksen KS, Andersen BB, et al. Detection of subclinical epileptiform discharges in Alzheimer’s disease using long-term outpatient EEG monitoring. Neurobiol. Dis. 2023;183:106149.

11. Devulder A, Macea J, Kalkanis A, et al. Subclinical epileptiform activity and sleep disturbances in Alzheimer’s disease. Brain Behav. 2023;13(12):e3306.

12. Vossel K, Ranasinghe KG, Beagle AJ, et al. Effect of Levetiracetam on Cognition in Patients With Alzheimer Disease With and Without Epileptiform Activity: A Randomized Clinical Trial. JAMA Neurol. 2021;78(11):1345.

13. Bakker A, Krauss GL, Albert MS, et al. Reduction of Hippocampal Hyperactivity Improves Cognition in Amnestic Mild Cognitive Impairment. Neuron 2012;74(3):467–474.

14. Kazim SF, Seo JH, Bianchi R, et al. Neuronal Network Excitability in Alzheimer’s Disease: The Puzzle of Similar versus Divergent Roles of Amyloid β and Tau. eneuro 2021;8(2):ENEURO.0418-20.2020.

15. Brunetti V, D’Atri A, Della Marca G, et al. Subclinical epileptiform activity during sleep in Alzheimer’s disease and mild cognitive impairment. Clin. Neurophysiol. 2020;131(5):1011–1018.

16. B. Szabo A, Cretin B, Gérard F, et al. Sleep: The Tip of the Iceberg in the Bidirectional Link Between Alzheimer’s Disease and Epilepsy. Front. Neurol. 2022;13:836292.

17. B. Szabo A, Cattaud V, Bezzina C, et al. Neuronal hyperexcitability in the Tg2576 mouse model of Alzheimer’s disease – the influence of sleep and noradrenergic transmission. Neurobiol. Aging 2023;123:35–48.

18. Kam K, Duffy ÁM, Moretto J, et al. Interictal spikes during sleep are an early defect in the Tg2576 mouse model of β-amyloid neuropathology. Sci. Rep. 2016;6(1):20119.

19. Frauscher B, Von Ellenrieder N, Dubeau F, Gotman J. EEG desynchronization during phasic REM sleep suppresses interictal epileptic activity in humans. Epilepsia 2016;57(6):879–888.

20. Shouse MN, Farber PR, Staba RJ. Physiological basis: how NREM sleep components can promote and REM sleep components can suppress seizure discharge propagation. Clin. Neurophysiol. 2000;111:S9–S18.

21. Ho A, Hannan S, Thomas J, et al. Rapid eye movement sleep affects interictal epileptic activity differently in mesiotemporal and neocortical areas. Epilepsia 2023;64(11):3036–3048.

22. Duffy ÁM, Morales-Corraliza J, Bermudez-Hernandez KM, et al. Entorhinal cortical defects in Tg2576 mice are present as early as 2–4 months of age. Neurobiol. Aging 2015;36(1):134–148.

23. Montplaisir J, Petit D, Gauthier S, et al. Sleep disturbances and eeg slowing in alzheimer’s disease. Sleep Res. Online 1998;1(4):147–151.

24. Petit D, Montplaisir J, Lorrain D, Gauthier S. Spectral analysis of the rapid eye movement sleep electroencephalogram in right and left temporal regions: A biological marker of Alzheimer’s disease. Ann. Neurol. 1992;32(2):172–176.

25. Puranen A, Taipale H, Koponen M, et al. Incidence of antidepressant use in community-dwelling persons with and without Alzheimer’s disease: 13-year follow-up. Int. J. Geriatr. Psychiatry 2017;32(1):94–101.

26. Slater IH, Jones GT, Moore RA. Inhibition of REM sleep by fluoxetine, a specific inhibitor of serotonin uptake. Neuropharmacology 1978;17(6):383–389.

27. Wichniak A, Wierzbicka A, Walęcka M, Jernajczyk W. Effects of Antidepressants on Sleep. Curr. Psychiatry Rep. 2017;19(9):63.

28. Stewart D, Johnson EL. The Bidirectional Relationship Between Epilepsy and Alzheimer’s Disease. Curr. Neurol. Neurosci. Rep. 2025;25(1):18.

29. Horvath A, Kiss M, Szucs A, Kamondi A. Precuneus-Dominant Degeneration of Parietal Lobe Is at Risk of Epilepsy in Mild Alzheimer’s Disease [Internet]. Front. Neurol. 2019;10[cited 2025 Aug 6 ] Available from: https://www.frontiersin.org/journals/neurology/articles/10.3389/fneur.2019.00878/full

30. Kane N, Acharya J, Beniczky S, et al. A revised glossary of terms most commonly used by clinical electroencephalographers and updated proposal for the report format of the EEG findings. Revision 2017. Clin. Neurophysiol. Pract. 2017;2:170–185.

31. Berry RB, Brooks R, Gamaldo C, et al. AASM Scoring Manual Updates for 2017 (Version 2.4). J. Clin. Sleep Med. 2017;13(05):665–666.

32. Fischl B, Salat DH, Van Der Kouwe AJW, et al. Sequence-independent segmentation of magnetic resonance images. NeuroImage 2004;23:S69–S84.

33. Fischl B, Salat DH, Busa E, et al. Whole Brain Segmentation. Neuron 2002;33(3):341–355.

34. Fischl B, Dale AM. Measuring the thickness of the human cerebral cortex from magnetic resonance images. Proc. Natl. Acad. Sci. 2000;97(20):11050–11055.

35. Yrondi A, Nemmi F, Billoux S, et al. Grey Matter changes in treatment-resistant depression during electroconvulsive therapy. J. Affect. Disord. 2019;258:42–49.

36. Kaestner E, Reyes A, Chen A, et al. Atrophy and cognitive profiles in older adults with temporal lobe epilepsy are similar to mild cognitive impairment. Brain 2021;144(1):236–250.

37. Love J, Selker R, Marsman M, et al. JASP : Graphical Statistical Software for Common Statistical Designs [Internet]. J. Stat. Softw. 2019;88(2)[cited 2024 Apr 17] Available from: http://www.jstatsoft.org/v88/i02/

38. Frauscher B, von Ellenrieder N, Ferrari-Marinho T, et al. Facilitation of epileptic activity during sleep is mediated by high amplitude slow waves. Brain 2015;138(6):1629–1641.

39. Romanelli MF, Morris JC, Ashkin K, Coben LA. Advanced Alzheimer’s Disease Is a Risk Factor for Late-Onset Seizures. Arch. Neurol. 1990;47(8):847–850.

40. Chu C-S, Lee F-L, Bai Y-M, et al. Antidepressant drugs use and epilepsy risk: A nationwide nested case-control study. Epilepsy Behav. 2023;140:109102.

41. Tallarico M, Pisano M, Leo A, et al. Antidepressant Drugs for Seizures and Epilepsy: Where do we Stand? Curr. Neuropharmacol. 2023;21(8):1691–1713.

42. Josephson CB, Lowerison M, Vallerand I, et al. Association of Depression and Treated Depression With Epilepsy and Seizure Outcomes: A Multicohort Analysis. JAMA Neurol. 2017;74(5):533–539.

43. Sierksma ASR, De Nijs L, Hoogland G, et al. Fluoxetine Treatment Induces Seizure Behavior and Premature Death in APPswe/PS1dE9 Mice. J. Alzheimers Dis. 2016;51(3):677–682.

44. David R, Manera V, Fabre R, et al. Evolution of the Antidepressant Prescribing in Alzheimer’s Disease and Related Disorders Between 2010 and 2014: Results from the French National Database on Alzheimer’s Disease (BNA). J. Alzheimers Dis. 2016;53(4):1365–1373.

45. Lam AD, Deck G, Goldman A, et al. Silent hippocampal seizures and spikes identified by foramen ovale electrodes in Alzheimer’s disease. Nat. Med. 2017;23(6):678–680.

46. Ranasinghe KG, Kudo K, Hinkley L, et al. Neuronal synchrony abnormalities associated with subclinical epileptiform activity in early-onset Alzheimer’s disease. Brain 2022;145(2):744–753.

47. Abou Jaoude M, Jacobs CS, Sarkis RA, et al. Noninvasive Detection of Hippocampal Epileptiform Activity on Scalp Electroencephalogram. JAMA Neurol. 2022;79(6):614–622.

48. Martínez-Cañada P, Perez-Valero E, Minguillon J, et al. Combining aperiodic 1/f slopes and brain simulation: An EEG/MEG proxy marker of excitation/inhibition imbalance in Alzheimer’s disease. Alzheimers Dement. Diagn. Assess. Dis. Monit. 2023;15(3):e12477.

49. Brookes MJ, Leggett J, Rea M, et al. Magnetoencephalography with optically pumped magnetometers (OPM-MEG): the next generation of functional neuroimaging. Trends Neurosci. 2022;45(8):621–634.

50. Horvath A, Kiss M, Szucs A, Kamondi A. Precuneus-Dominant Degeneration of Parietal Lobe Is at Risk of Epilepsy in Mild Alzheimer’s Disease. Front. Neurol. 2019;10:878.

51. Shokri-Kojori E, Wang G-J, Wiers CE, et al. β-Amyloid accumulation in the human brain after one night of sleep deprivation. Proc. Natl. Acad. Sci. 2018;115(17):4483–4488.

52. André C, Rehel S, Kuhn E, et al. Association of Sleep-Disordered Breathing With Alzheimer Disease Biomarkers in Community-Dwelling Older Adults: A Secondary Analysis of a Randomized Clinical Trial. JAMA Neurol. 2020;77(6):716–724.

53. Busche MA, Chen X, Henning HA, et al. Critical role of soluble amyloid-β for early hippocampal hyperactivity in a mouse model of Alzheimer’s disease. Proc. Natl. Acad. Sci. 2012;109(22):8740–8745.

54. Casula EP, Borghi I, Maiella M, et al. Regional Precuneus Cortical Hyperexcitability in Alzheimer’s Disease Patients. Ann. Neurol. 2023;93(2):371–383.

55. Pasquini L, Rahmani F, Maleki-Balajoo S, et al. Medial Temporal Lobe Disconnection and Hyperexcitability Across Alzheimer’s Disease Stages. J. Alzheimers Dis. Rep. 2019;3(1):103–112.

## References

B. Szabo, A., Cretin, B., Gérard, F., Curot, J., J. Barbeau, E., Pariente, J., Dahan, L. & Valton, L. (2022). Sleep: The Tip of the Iceberg in the Bidirectional Link Between Alzheimer’s Disease and Epilepsy. Frontiers in Neurology, 13, 836292.

